# Pubertal growth spurt is not universal: Polymorphism in growth at adolescence and its possible origin

**DOI:** 10.1101/2023.03.24.23287682

**Authors:** Maciej Henneberg, Iwona Rosset, Elżbieta Żądzińska

**Author notes:** **Corresponding Author:** Maciej Henneberg, School of Biomedicine. The University of Adelaide. Adelaide. South Australia 5005. Australia. Maciej. MH, IR, EŻ contributed equally to this paper. **Contributor and guarantor information** MH and EŻ conceived the hypothesis, IR and EŻ obtained data, MH and IR conducted statistical analyses and all three authors interpreted results and edited the text. The corresponding author attests that all listed authors meet authorship criteria and that no others meeting the criteria have been omitted. **Copyright licence** The Corresponding Author has the right to grant on behalf of all authors and does grant on behalf of all authors, a worldwide licence to the Publishers and its licensees in perpetuity, in all forms, formats and media (whether known now or created in the future), to i) publish, reproduce, distribute, display and store the Contribution, ii) translate the Contribution into other languages, create adaptations, reprints, include within collections and create summaries, extracts and/or, abstracts of the Contribution, iii) create any other derivative work(s) based on the Contribution, iv) to exploit all subsidiary rights in the Contribution, v) the inclusion of electronic links from the Contribution to third party material where-ever it may be located; and, vi) licence any third party to do any or all of the above. **Transparency declaration** The lead author* affirms that this manuscript is an honest, accurate, and transparent account of the study being reported; that no important aspects of the study have been omitted; and that any discrepancies from the study as planned (and, if relevant, registered) have been explained.

## Abstract

**Objectives:** Numerous studies of child growth conducted in industrialised countries showed presence of the phenomenon of the adolescent growth spurt to the extent that its presence became a dogma applied to humans as a species. However, earliest observations of growth at adolescence and also observations of adolescent growth conducted in small traditional societies do not show pubertal spurts consistently. Longitudinal observations of growth of individuals in present-day societies show strong polymorphism of the age at which puberty starts and ends and the magnitude of growth acceleration during puberty. Some normally developing individuals may not experience the pubertal growth spurt.

**Design, Seeting, Participants:** Longitudinal height growth data of 110 girls aged 6-18 years from homogenous socio-economic situation – one medical high school (=nursing college) in one large Polish city of Lodz.

**Results:** 18 (16%) girls did not show statistically significant acceleration of body height growth while they reached the same adult height as their spurting peers. Girls who experienced growth spurt had the age at peak height velocity of 11.2 years and peak height velocity of 69.3 mm, comparable to those found in many other studies. There was a negative correlation of adult height with variation of individual accelerations (r-0.24, p=0.01) – girls whose accelerations differed less from year to year achieved greater adult heights.

**Conclusions:** Findings of no pubertal spurts in some individuals have been made in other samples studied by other authors, though rarely reported. It can be argued that slow growth at adolescence was more favourable in conditions of limited access to nutrition and medical care while polymorphisms determining fast, uneven growth became more prevalent after industrialisation that, together with progress in health sciences, relaxed natural selection on patterns of growth. Individuals who do not experience pubertal growth spurts, but are otherwise healthy, should not be subject to clinical interventions.

**Summary Box:** *What is already known on this topic:* Pubertal growth spurt in body height is considered a typical feature of postnatal development. There is a large polymorphism of the age at which the spurt occurs and of its intensity. Individuals who do not experience the spurt, but are otherwise healthy, may be subject to uncesessary clinical interventions.

*What this study adds:* Among 110 female students of an urban secondary nursing college studied longitudinally from 6 to 16 years, 16% did not experience acceleration of growth at puberty. Similar findings were reported in the literature from some non-Westernised communities and from European 19^th^ century populations.

## Introduction

Human postnatal ontogeny got extended during evolution of hominins by the insertion of childhood, a phase of relatively slow growth of sexually immature individual. [1, 2] This gives time to learn complex social and technological skills required for life in the human society. The growth rate after being fast in the period of infancy slows down for several years during which children have time to learn while they elicit caring behaviours of adults by being small and asexual. Eventually, delayed sexual maturation occurs. During this period changes in hormonal body regulation accelerate further growth in size to reach adult body dimensions Tanner. [3] This growth occurs often at rates faster than in childhood and is described as the pubertal growth spurt. Pubertal changes resemble, to a certain extent, a biological metamorphosis that occurs in the development of some invertebrates and vertebrates (e.g. amphibians). Avereage body weights of some primates show changes with age that can be considered pubertal growth spurts. [4]

It has been noticed in observations of single individuals as early as the 18^th^ century (Francois de Montbeillard) while during the 20^th^ century this observation resulted in intensive and extensive research aimed at measuring its most interesting characteristics: the peak height velocity of growth, its age in relation to signs of sexual maturation (especially menarche in females) and its course – the onset, duration and end. Influence of environmental and genetic factors on these characteristics were studied with the conclusion that genetic variation plays a significant role in differentiating individual growth. [5, 6] Already early on a great variability of age at which characteristics of sexual maturation occur and similarly large variability of the magnitude of growth and its acceleration were noticed and documented. Burk in 1898 summarised findings of many 19^th^ century American and European studies as follows:

“*Individual children of normal rate of growth show a comparatively wide range of deviation at any given period, and also a wide deviation in reaching the crises in the periods of growth*. “. [7]

This variability has been thoroughly documented in the early 20^th^ century leading Tanner et al. in construction of the whole year velocity standards of British children to the conclusion that “At adolescence these standards are greatly scattered by the phase-difference effect.” [8, 9, 10]

During the spurt musculoskeletal system grows very fast, so fast in fact that neuromuscular coordination may become impaired, physiological homeostasis is disturbed producing cardiovascular inadequacies and excessive caloric requirements. Neurohormonal changes result in behavioural imbalance. The fast production of new quantities of tissues increases nutritional demands beyond the adult caloric needs. Spurting adolescents are energetically expensive, behaviourally and physically imbalanced individuals potentially exposed to dangers of environmental and social maladaptation. [11, 12, 13]

It seems therefore that the adolescent growth spurt is maladaptive, especially in the context of living conditions of past populations with limited food resources and poor health care. Unfortunately, the vast majority of large growth studies was conducted in developed industrial nations in the last two centuries. There is no information on how large and significant adolescent growth spurt occurred before the modern times. The early systematic studies of child growth in European population samples did not document a substantial increase in growth rates at adolescence. [14] Alphonse Quetelet (1831) tried to establish “the rule of human growth” (*la loi de croissance de l’homme)* where he assumed that the growth velocity decreases monotonically from infancy to adulthood. [15] He stated clearly (p. 21): “*A partir de 4 à 5 ans, l’accroissement de taille deviant a peu pres exactement regulier jusque vers 16 ans, c’est-à-dire jusqu’après l’âge de la puberté*’’. He based this on cross-sectional data collected in Belgium, however, the data he published indicate a slight increase of growth rate at adolescence (Fig. 1). Earlier, longitudinal observation of growth of one boy – Francois de Montbeillard published by Georges-Louis Leclerc de Buffon in his 1777 supplement to l’Historie Naturelle de l’Homme indicated an increased growth velocity at adolescence. However, this son of a wealthy family became unusually tall (1,868 mm) at the age 17.6 years as compared with 1,640 mm of 17 years old Belgian boys in early 19^th^ century. [15] The difference of 228 mm exceeds three times (3.3-3.5) a standard deviation of body height of boys at this age that is approximately 65-70 mm. Values of growth velocities calculated from the Buffon’s data table of Francois’s heights vary irregularly showing, however, some acceleration at approximately 12 to 15 years (Fig. 2). By the end of the 19^th^ century, summaries of studies of growth of large samples of children, using principles of essentialism, that is concentrating on statistical measures of central tendency, established that at puberty an increase in the rate of growth in height is standard, a basic characteristics of human growth pattern. [7] This has been widely accepted.

**Fig. 1.**
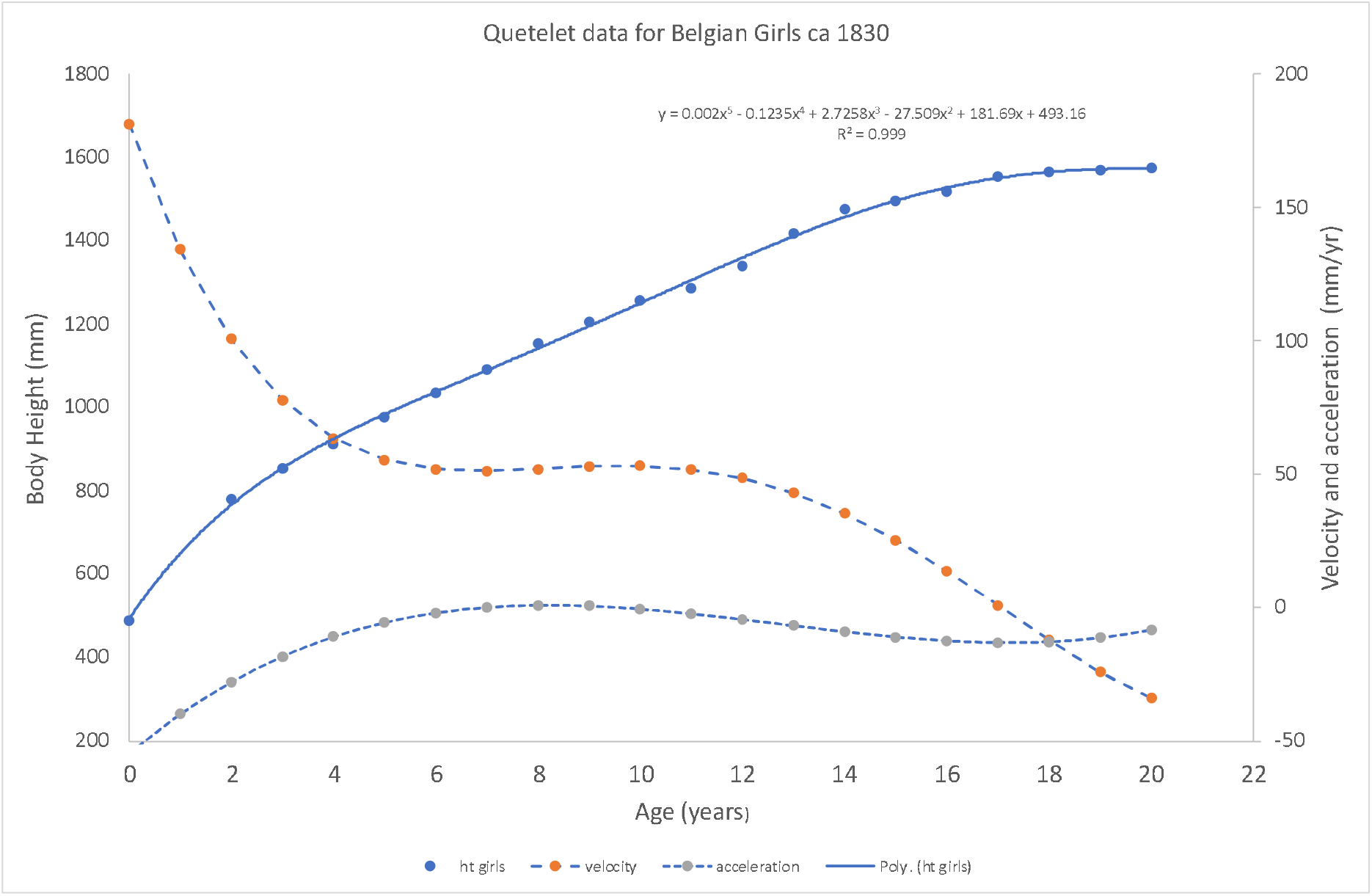
Quetelet’s data for Belgian girls ca 1830. Polynomial regression was fitted to the original data and then velocity and acceleration calculated as the first and the second derivative of the polynomial regression equation. Note the lack of obvious increase in velocity at puberty and corresponding near-zero values of acceleration.

**Fig. 2.**
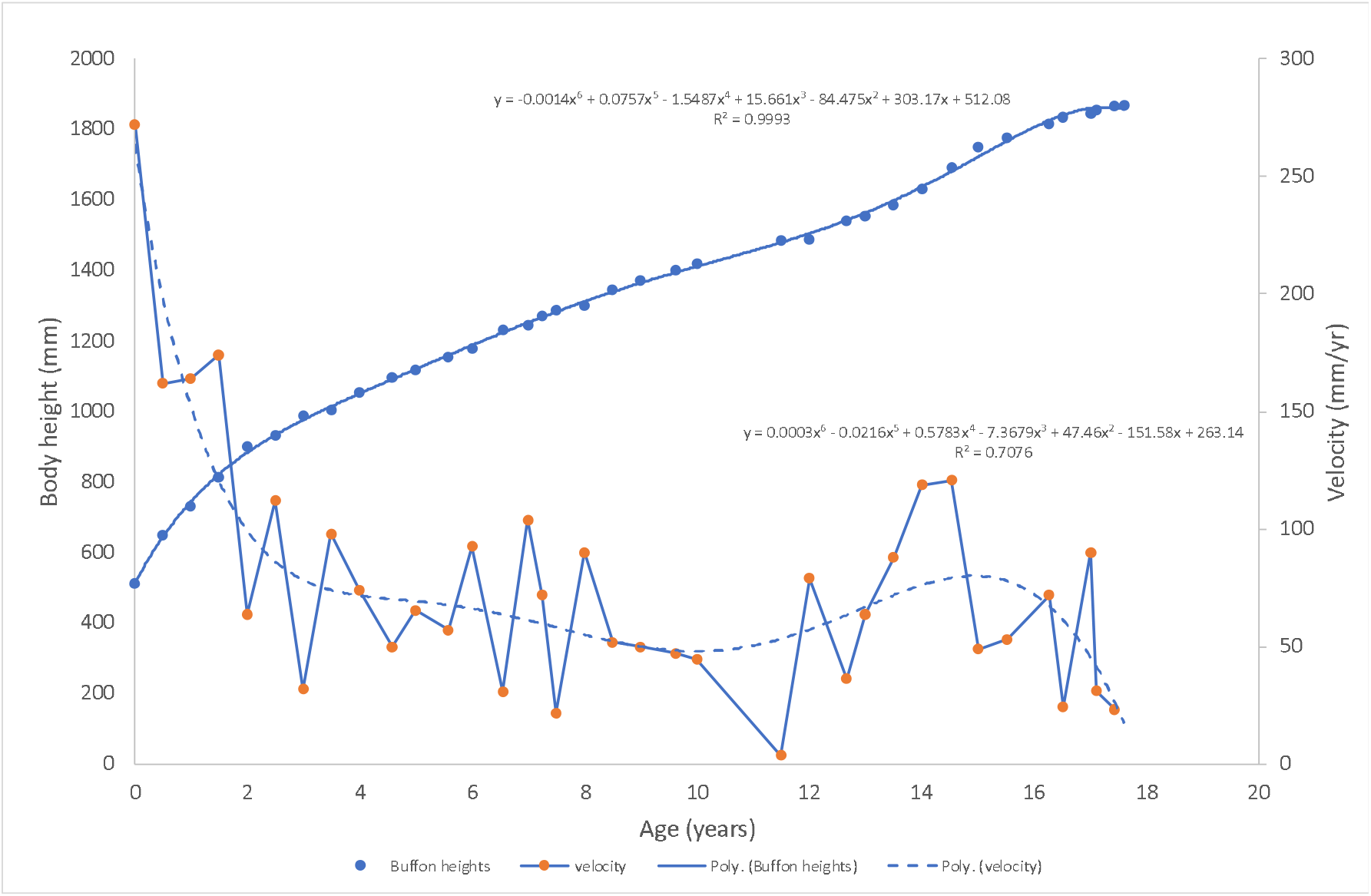
Buffon’s longitudinal data for Francois de Montbeillard’s growth in body height. Velocities are calculated as annual ones by subtracting the previous measurement from the next and dividing by a fraction of a year that elapsed between the consecutive measurements.

Recent studies of child growth in the 19^th^ century did not find the growth spurt in children of lower socioeconomic status. [16] Similarly, many individual children in an African rural community did not experience significant acceleration of their growth rates at adolescence. [17] Walker et al. did not find adolescent growth spurt in a number of “small-scale societies” with traditional, non-Western lifestyles. [18] As suggested by Budnik and Henneberg “Social mobility to higher classes [in the 19^th^ century] produced the phenomenon of growth spurt.” [16]

Biological variation is a fundamental property of living systems. Growth rates and their age profiles are variable among human individuals. Ontogenetic events are under strong genetic control. [19] Even in the same socioeconomic conditions some children grow faster, some slower and rates of their growth do not occur with the same magnitude at same ages. It may be that spectacular adolescent growth spurts that attracted attention of modern researchers occur only in some individuals that appeared in larger numbers only when living conditions became permissive to maladaptive phenomena of increased caloric requirements, impaired neuromotor coordination and behavioural inadequacies. Growth of some individuals without pubertal spurts seems to be quite normal as it is an expression of normal biological variation.

We hypothesise that adolescent growth spurts are an expression of heritable biological variation in ontogeny in good living conditions, not a standard common phenomenon, and thus they are not present in all normally developing individuals.

## Participants and Method

Material for this study was collected as a part of a research program monitoring the development of preschool and school children in the years 1992-1994. [20] Measurements of children were carried out in Łódź (a city of 700,000 people located in central Poland) and studied according to the procedure used recently by Henneberg & żądzińska. [21]

The research was carried out in collaboration with the local education administration (Board of Education), school heads, parents and children. Each child was asked for oral consent to be examined by a member of the anthropometric team, and children who refused were not measured. All members of the research crew spoke the local language (Polish). The whole procedure was approved by the Research Bioethics Committee of the University of Lodz (KBBN-UŁ).

Retrospective data were collected from health cards of 388 students of the Medical High School No. 1 in Łódź, belonging to the Medical School Complex (122 Narutowicza Street, 90-145 Łódź). Health records contained information collected during preventive medical examinations carried out by primary care doctors and nurses (procedure according to the rules currently in force in Poland: https://nfz-gdansk.pl/uploads/files/Dz_U_2019_736.pdf). Here we use annual measurements of body height of girls born in 1973-1976, living in Łódź and neighbouring towns and villages. Decimal age of each female at each measurement occasion was calculated to the nearest one hundredth from the date of birth and dates of primary medical care visits. Data selected for this study included females for whom the first measurement was taken at age lower than 6.51 years and the last one at age exceeding 15.50 years and who were measured regularly at consecutive intervals of approximately one year or less in the age range 9.00 – 15.00 years. This selection resulted in 110 longitudinal records analyzed here (Supplementary information).

Measurement error and reliability of measurements were not determined at the time of data collection. However, girls were measured annually until they left the school at age 16-19 years. The last few measurements of many individuals did not increase regularly, but just fluctuated randomly. Since females may attain their final adult stature at the age of 16 years repeated measurements after that age will differ only because of random errors of measurement. Altogether 76 participants did not show regular body height increases in the last few years of their participation in the study. Their consecutive measurements were treated as test-retest pairs and the Technical Error of Measurement was calculated following the formula TEM = SQR(((x_1_-x_2_)^2)/2N). It amounted to 6.4 mm. Reliability, that is a coefficient of correlation between the first and the second measurement was 0.9851.

Numerous methods for studying the growth at adolescence have been developed. The automatic assumption of the pubertal growth acceleration and thus growth peaks, lies at the basis of a number of them. [22, 23, 24, 25, 26, 27] Application of these methods would lead to circular reasoning in our research. Other methods use various polynomials that are theoretically assumption-free but differ in details of their applications. [28, 29, 30, 31, 32] Various methods produce different results concerning age at peak height velocity, peak height velocity and other characteristics, when applied to the same data. [30, 31] To avoid forcing assumptions into our data set we have chosen an assumption-free method of fitting polynomial regressions to longitudinal growth data.

Individual longitudinal records of body height, were fitted by polynomial regressions. Their first derivatives were calculated as measures of growth velocities and second derivatives as measures of growth acceleration. Rather than fitting polynomials of a defined degree, for instance fifth degree polynomials as recommended by Boeyer et al., [32] when fitting a record of a particular individual, we kept increasing a degree of a polynomial from the second up until the goodness of fit at least equalled the reliability of measurements of body height (0.997). Any further increase of the polynomial’s degree would reflect random errors of measurement rather than biological reality. The presence of the growth spurt was assessed by establishing that the following year’s velocity was significantly greater than the previous year’s velocity that is the difference of consecutive velocities exceeded twice TEMs of two annual measurements. [17]

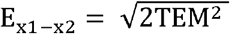

In the case of our material E_(x1-x2)_ = 9.05, thus any acceleration exceeding 18.1 mm/year^2^ can be considered significant. Lower values of acceleration may be a result of combination of measurement errors. This approach is correct when velocities are calculated by subtraction of actual values of annual measurements. Fitting of a polynomial regression line to all consecutive measurements of a given person acts like calculation of an arithmetic mean with regard to measurement errors. In this situation, error of a single measurement is divided by the number of measurements the regression line is fitted to. Therefore for an individual who had 10 measurements of body height to which a polynomial regression has been fitted, a significant acceleration obtained from the second derivative of the regression equation must exceed two times the error calculated from the TEM divided by 10. For example: SQR(2*(6.4/10)^2)=0.91. This criterion was applied to accelerations at ages exceeding 8.9 years. An individual was considered to have achieved a growth spurt when her acceleration, calculated as the second derivative of the polynomial regression, in the age range 9-15 years was significant by the above criterion. Variance of annual acceleration values for each girl was calculated as a measure of differences between annual accelerations. In those calculations data for the earliest and the latest age were not included because derivatives of polynomials tend to have unstable initial and final values. [32]

All calculations were made using Statistica data analysis software system version 13.3 and Microsoft Excel for Mac version 16.70.

## Results

Cross-sectional analysis of body heights by age (Table 1.) in our sample shows a pattern similar to that of other European girls.

**Table 1.** Cross-sectional body height of girls. Age of the last birthday. N=number of longitudinal observations at a given age

| Age | average | stdev | N |
| --- | --- | --- | --- |
| 5 | 32 | 1141.6 | 54.7 |
| 6 | 91 | 1184.5 | 46.8 |
| 7 | 102 | 1242.9 | 50.1 |
| 8 | 108 | 1295.2 | 53.0 |
| 9 | 113 | 1347.4 | 57.8 |
| 10 | 106 | 1401.0 | 62.7 |
| 11 | 110 | 1463.7 | 70.2 |
| 12 | 110 | 1531.9 | 65.9 |
| 13 | 113 | 1579.5 | 52.5 |
| 14 | 110 | 1610.7 | 50.5 |
| 15 | 134 | 1618.7 | 53.5 |
| 16 | 126 | 1629.8 | 49.9 |
| 17 | 37 | 1620.9 | 43.3 |
| 18 | 18 | 1640.8 | 47.6 |

**Table 2.** Characteristics of growth at adolescence of girls from the Medical High School in Łódź. Height values in millimetres, acceleration measured in milimetres per year squared. PHV – peak height velocity, APHV – age at peak height velocity, end ht – height attained (ie height that did not differ from heights measured at older ages), Age end ht – age at which the end height was attained.

|  | With significant spurt N=92 |  |  |  |  | without significant spurt N=18 |  |  |  |  |
| --- | --- | --- | --- | --- | --- | --- | --- | --- | --- | --- |
|  | PHV | APHV | Accel | end ht | Age end ht | PHV | APHV | Accel | end ht | Age end ht |
| Avg | 69.3 | 11.2<sup>1</sup> | 18.2<sup>2</sup> | 1631.7 | 16.3 | 66.3 | 9.7<sup>1</sup> | 9.5<sup>2</sup> | 1633.9 | 16.3 |
| SD | 12.0 | 1.1 | 15.0 | 48.9 | 1.2 | 9.6 | 1.8 | 5.8 | 56.4 | 0.8 |

<sup>1</sup> – difference between girls with spurt and without spurt is significant, p=0.0003, T=4.31
<sup>2</sup> - difference between girls with spurt and without spurt is significant, p=0.0002, T=4.09
(T-test for unequal variances)

We found that in the city of Łódź 92 girls had the pubertal spurt of body height, and 18 did not have a spurt. Figures 3-6 illustrate growth of some individuals differing in patterns of velocity and acceleration.

**Fig. 3.**
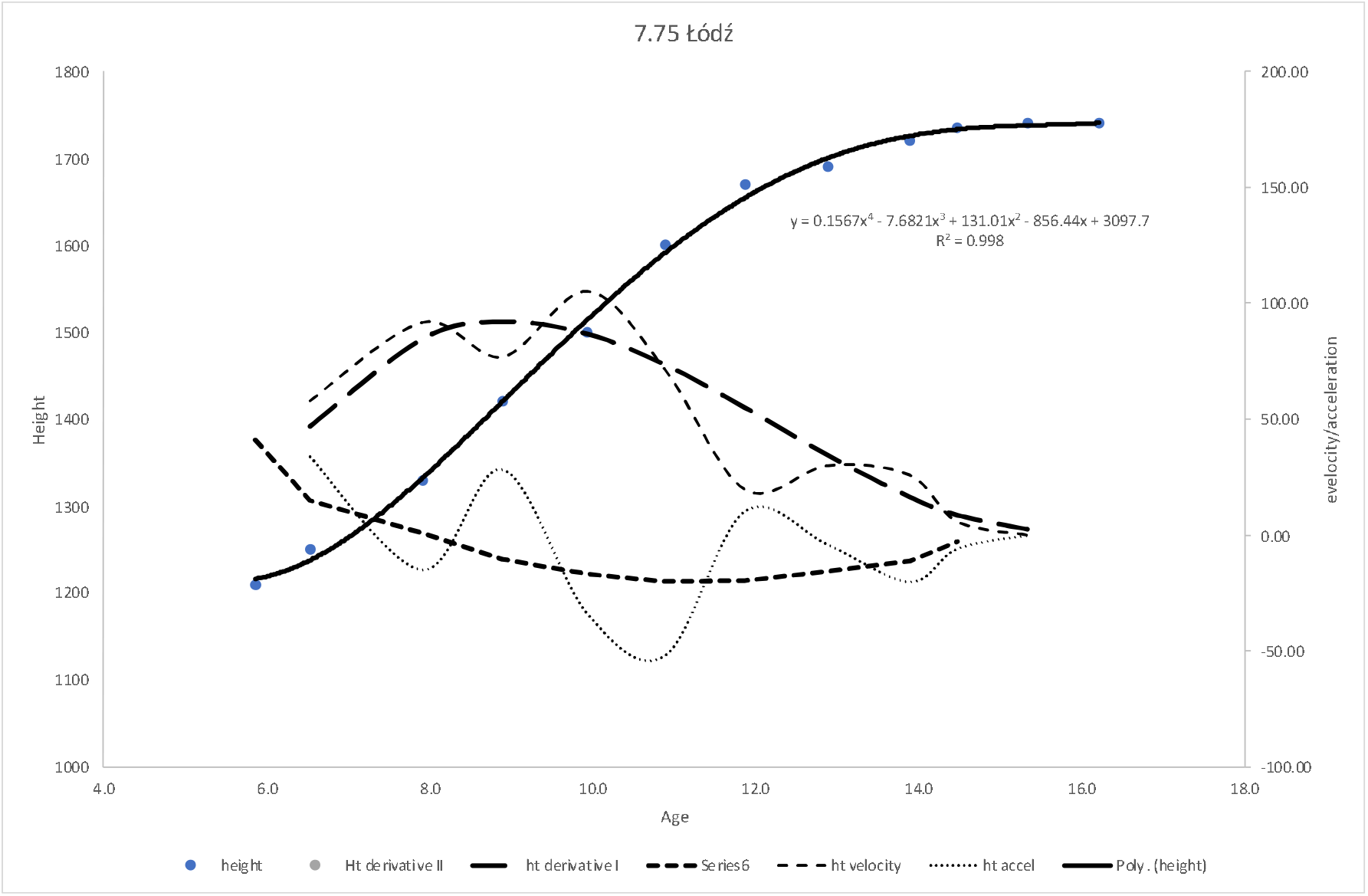
Polynomial regression fitted to the growth measurements of a girl #7.75. Velocity and acceleration were calculated in two ways: 1. As the ffirst and the second derivatives of the polynomial equation, 2 by subtraction of consecutive measurements and their standardisation for a full one year. These latter oones are influenced by measurement errors more than derivatives. Note the lack of abrupt change in acceleration during the puberty.

**Fig. 4.**
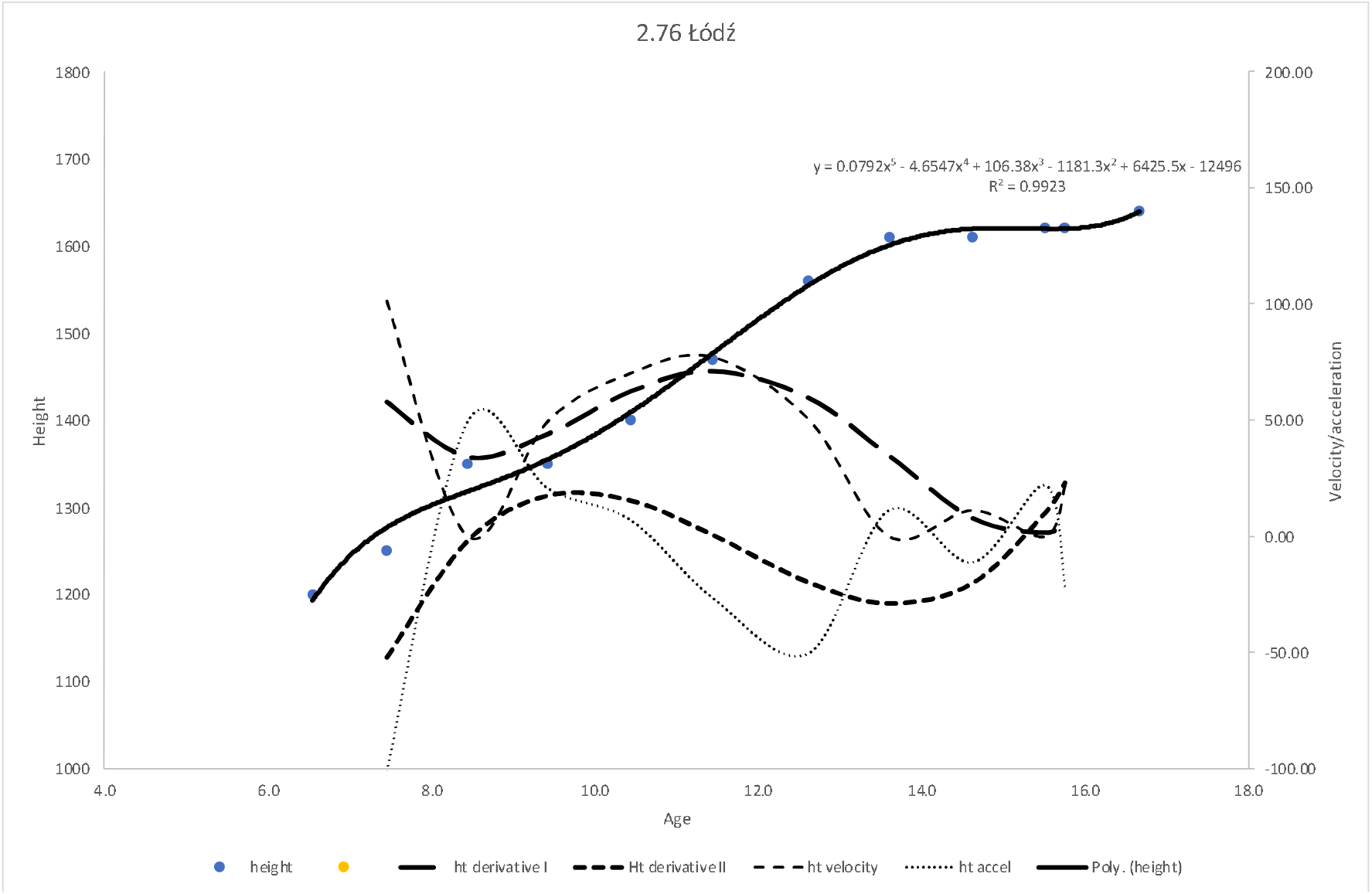
Polynomial regression fitted to the growth measurements of a girl #2.76. Velocity and acceleration were calculated in two ways: 1. As the first and the second derivatives of the polynomial equation, 2. by subtraction of consecutive measurements and their standardisation for a full one year. These latter ones are influenced by measurement errors more than derivatives. Note a strong increase of velocity aroundage 11 years and preceding increase of acceleration up to about 9-10 years.

**Fig. 5.**
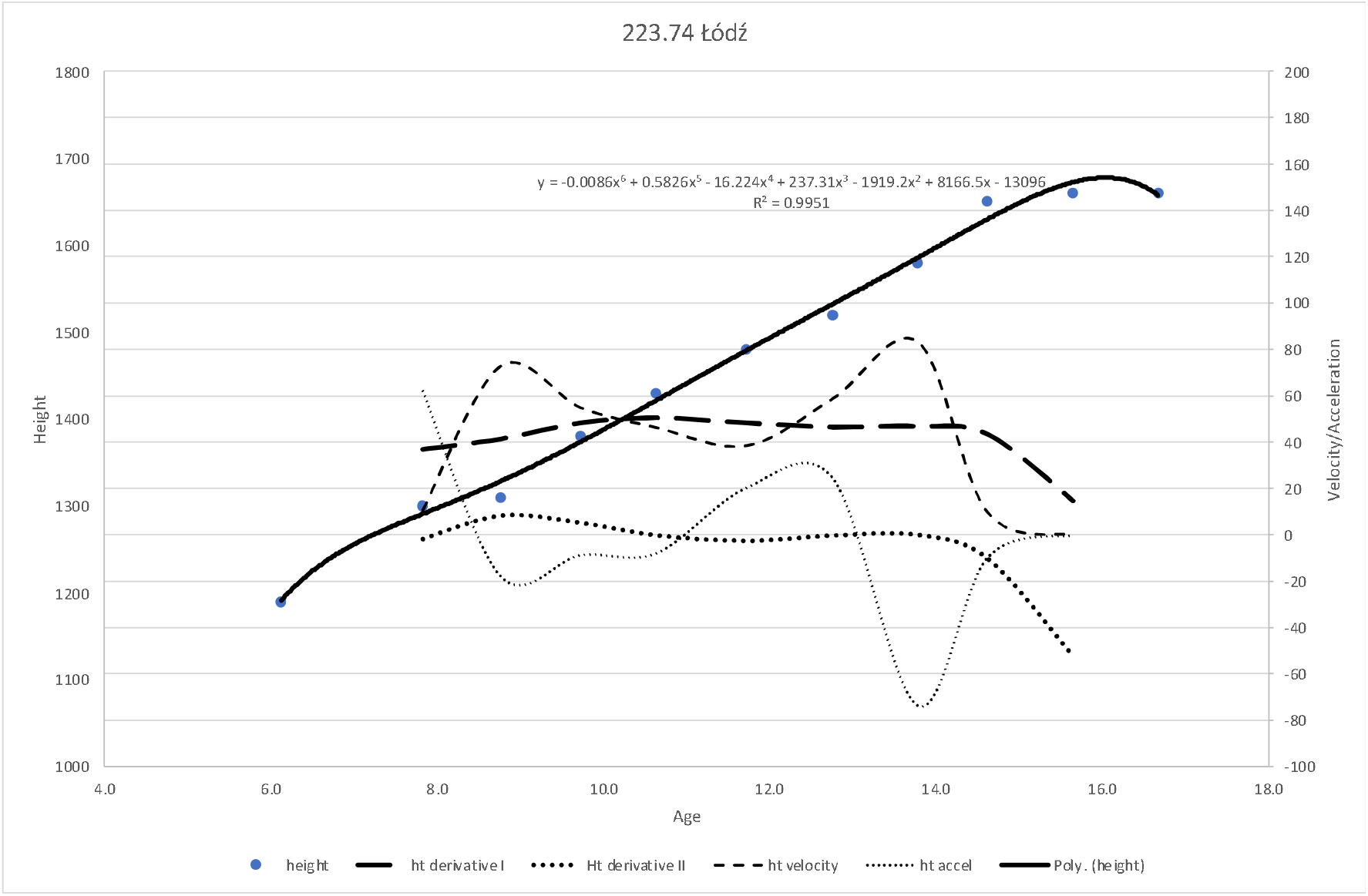
Polynomial regression fitted to the growth measurements of a girl #223.74. Velocity and acceleration were calculated in two ways: 1. As the first and the second derivatives of the polynomial equation, 2. by subtraction of consecutive measurements and their standardisation for a full one year. These latter ones are influenced by measurement errors more than derivatives.Note the lack of increased velocity from 10 to 14 years and corresponding near-zero acceleration.

**Fig. 6.**
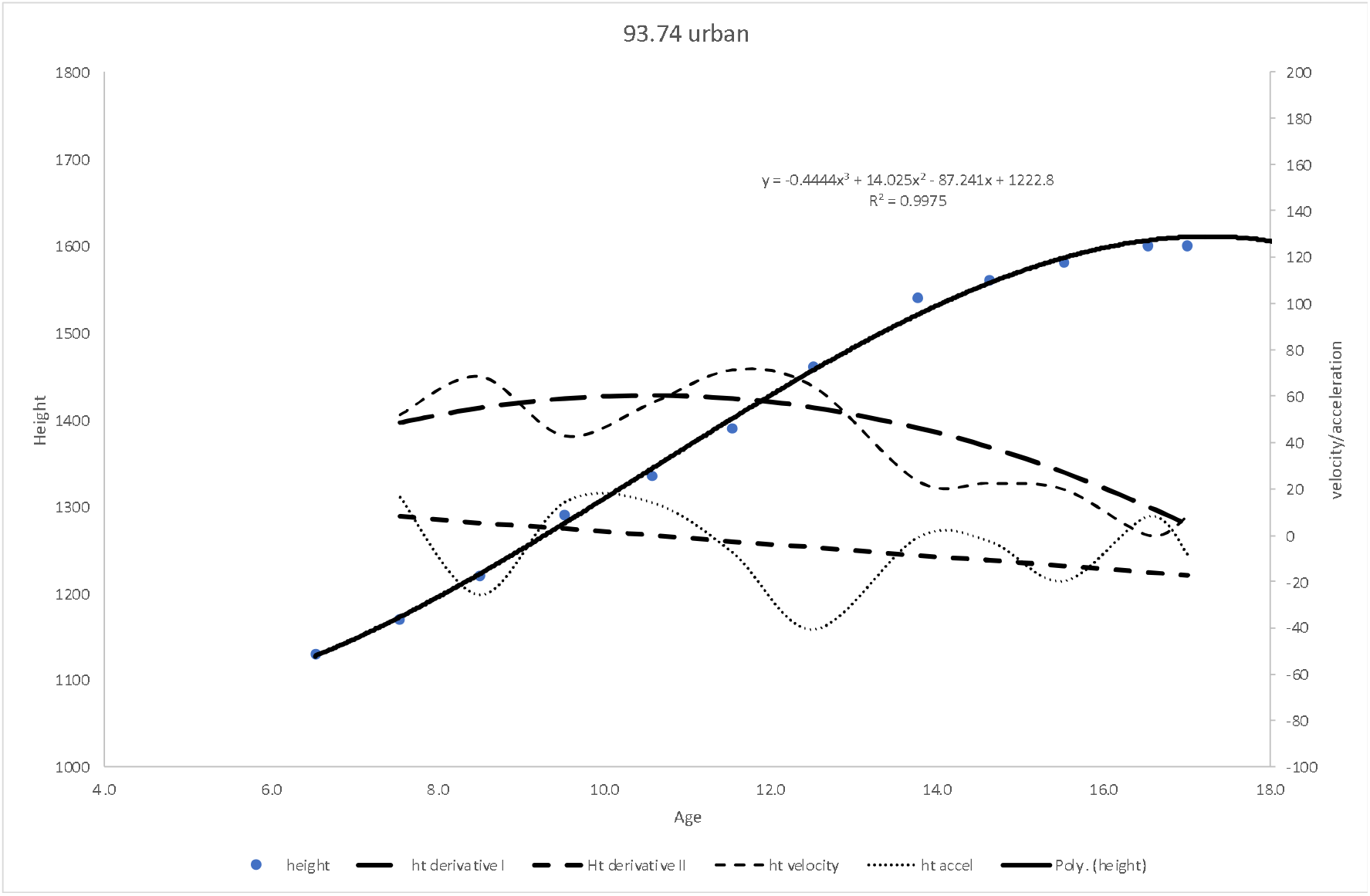
Polynomial regression fitted to the growth measurements of a girl #93.74. Velocity and acceleration were calculated in two ways: 1. As the first and the second derivatives of the polynomial equation, 2. by subtraction of consecutive measurements and their standardisation for a full one year. These latter ones are influenced by measurement errors more than derivatives. Note the lack of noticeable incease of velocity during puberty and a flat gently sloping down from near-zero line of aceleration.

Among 92 girls experiencing significant spurts 87 came from the urban living environment and five (5.4%) from the rural environment, while among 18 girls without significant growth spurt only one (5.6%) came from the rural environment, the remaining 17 grew up in the urban environment.

Individual variation of velocities and accelerations in both girls with significant spurt and without it is present, as expected in a polygenetically controlled trait subject to ecosensitive responses (Figures 7 and 8), but the distinction in magnitudes of age changes in both characteristics is clear. The average variance of accelerations in girls with the spurt is 743.1 (mm/year^2^)^2^ while it is only 235.4 (mm/year^2^)^2^ in girls without the spurt, more than 3 times less! Despite these differences there was no difference in adult height, girls without the spurt being slightly (insignificantly) taller. The same is true for average heights of these two groups of girls at the age 7 years (data not shown).

**Fig. 7.**
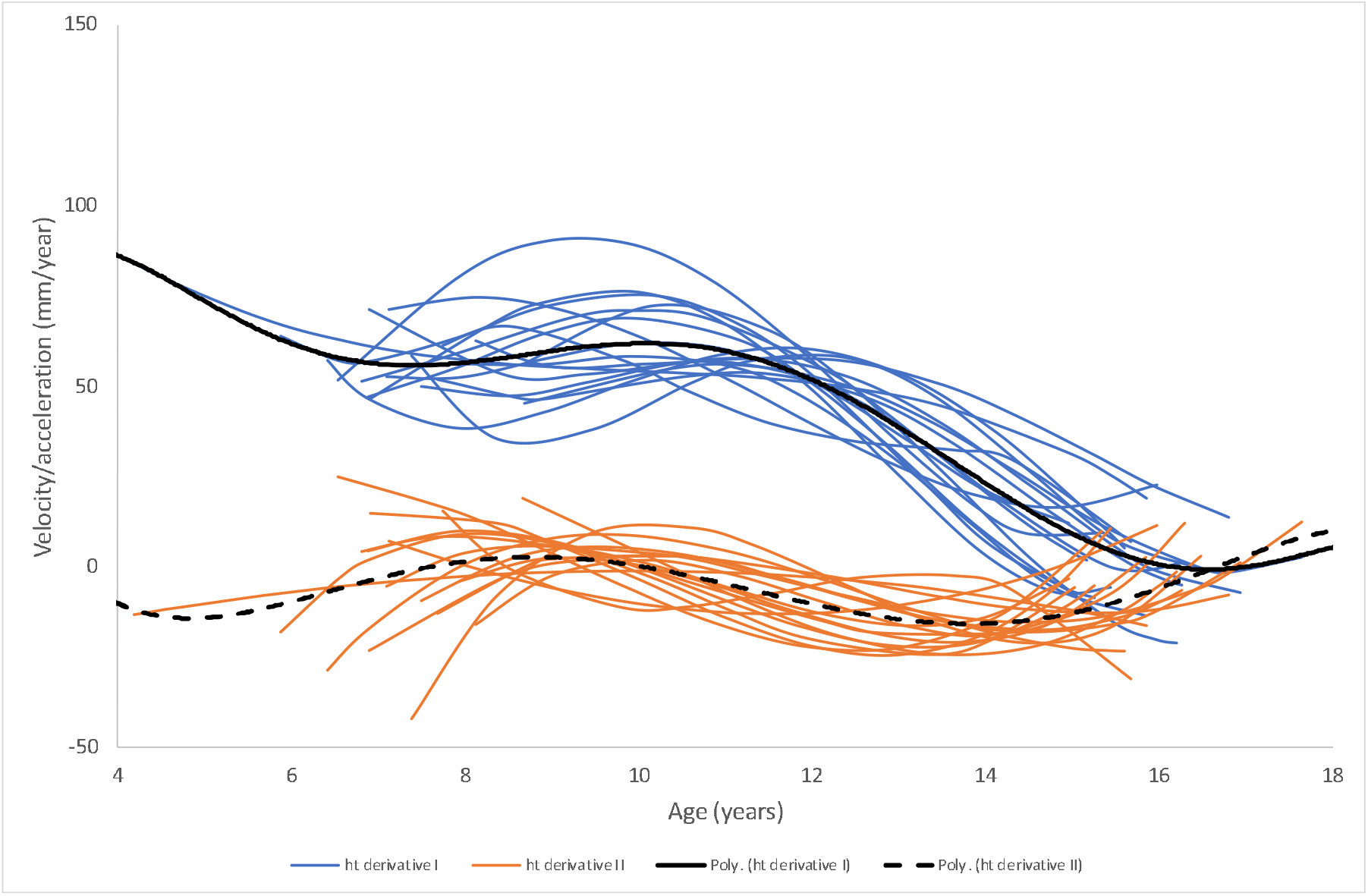
Females without a significant body height growth spurt. Velocity and acceleration are first and second derivatives of the fifth-degree polynomial fitted to their growth data.

**Fig. 8.**
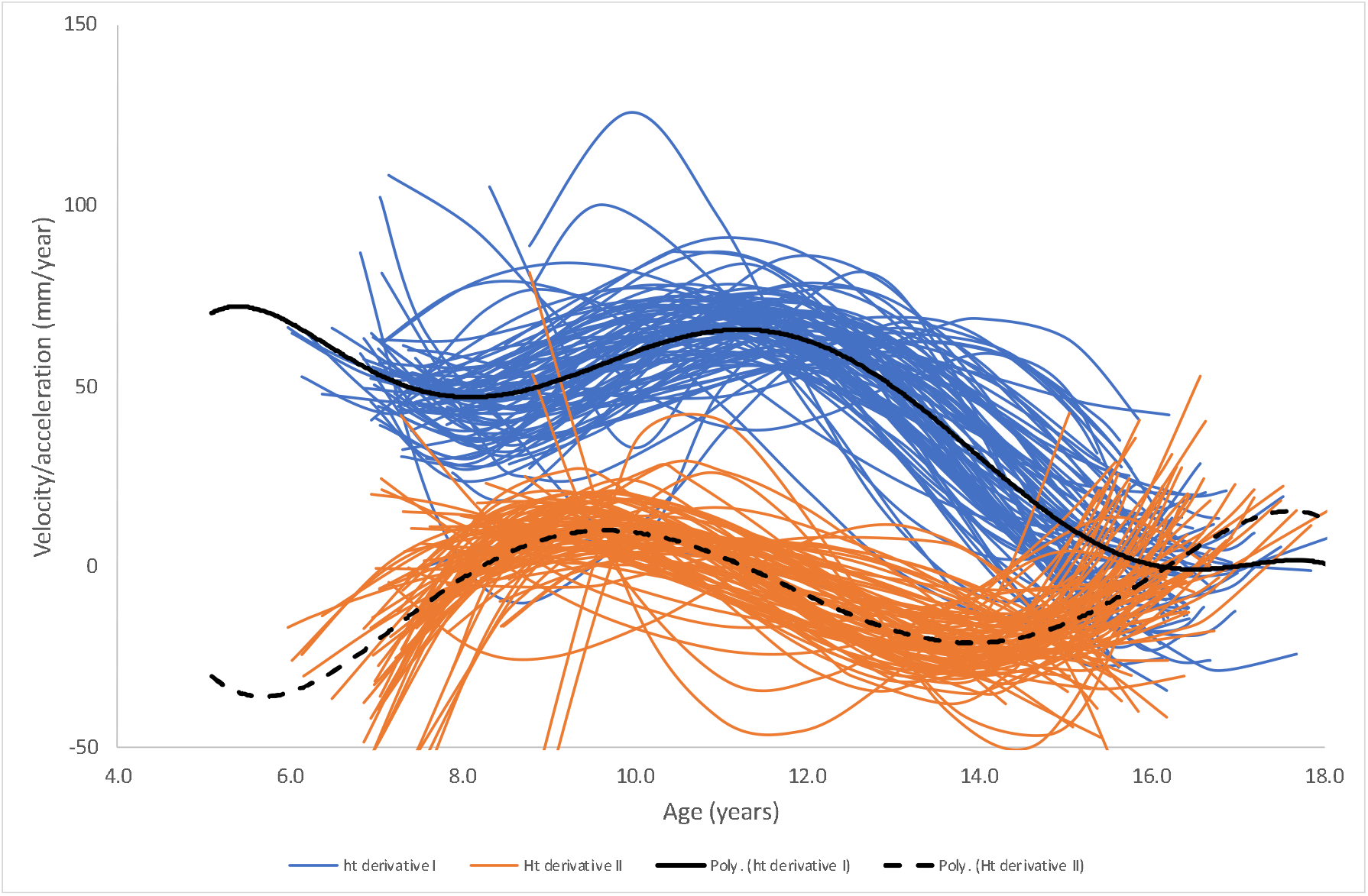
Females with significant body height growth spurts. Velocity and acceleration are first and second derivatives of the fifth-degree polynomial fitted to their growth data.

**Fig. 9.**
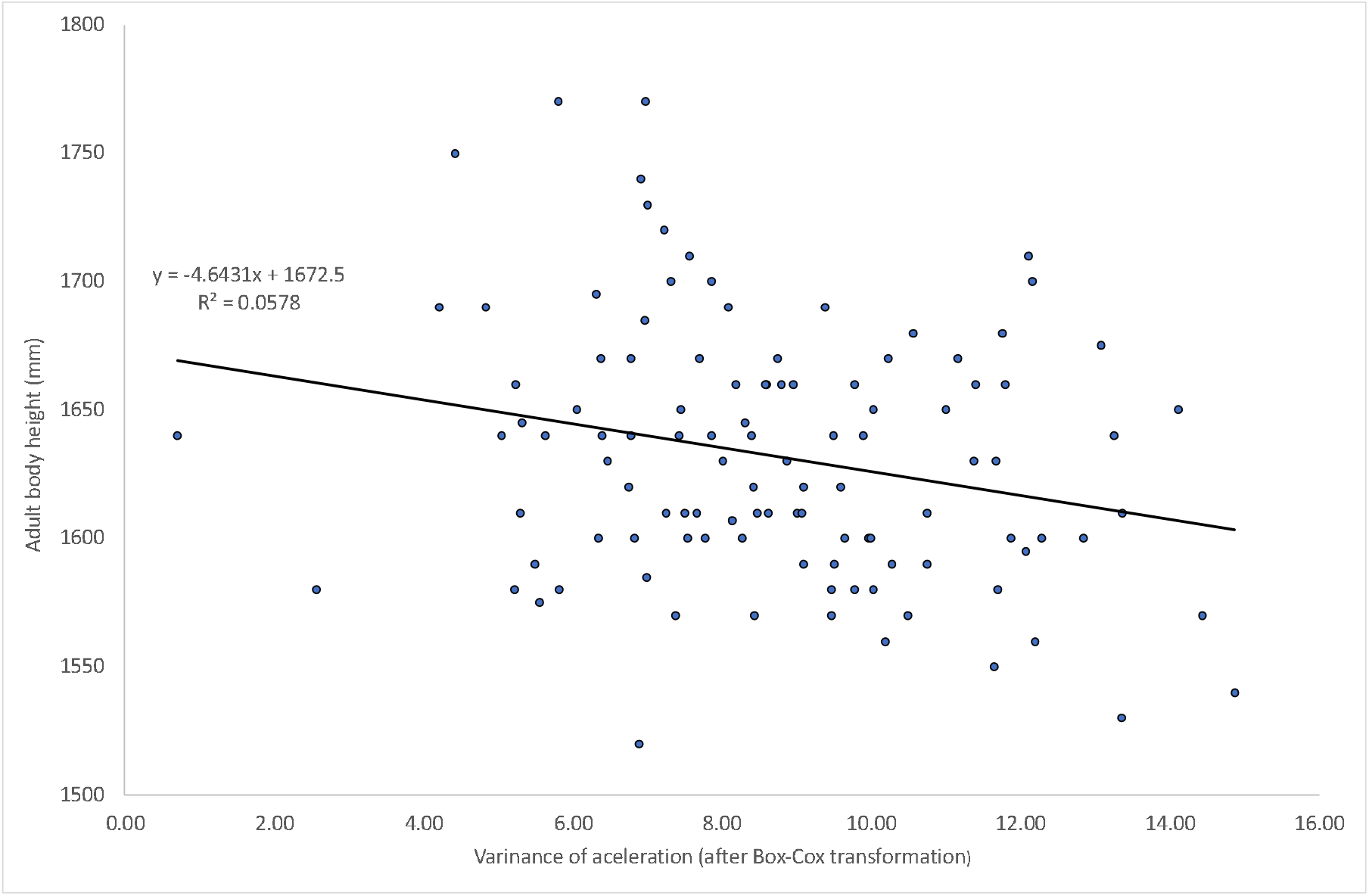
Regression of adult body height on the variance of individual acceleration during the puberty. Correlation (r=-0.2404) is significant, p=0.0114, N=110.

In the entire sample there is a negative correlation between adult body height and variances of velocities (r=-0.2404, p=0.0114, Fig.9). That is, the smaller are differences between annual accelerations, the taller height achieved at the end of growth period. The same correlation obtains when only girls with a spurt are observed (r=-0.3002, p= 0.0036). In the small sample (N=18) of the girls without spurt there is also similar negative correlation (r=-0.2243), but it does not reach the formal level of significance.

## Discussion

Peak height velocities and the age at peak height velocity in 92 girls with the spurt compare well with most of other populations studied, especially the classic studies of Tanner et al., [10] and Ramsay et al. [34] However, we have found that at least 16.4% of studied females had no significant adolescent growth spurt based on our error criteria favourable for detecting spurts, that is small magnitude of error. It compares with 40% of South African girls studied at the end of the 20^th^ century where criteria for spurt detection were less favourable, that is greater ranges of random error were used. [17]

It is important to note that girls who had no significant growth spurt attained the same adult height (even 2 mm greater) at exactly the same age as their “spurting” peers while they were also the same height as others at the beginning of the pubertal period. The individual variation of annual accelerations has a very wide range, what agrees well with the wide range of peak velocities and ages at peak height velocity. Clearly, in our sample there were different growth trajectories that arrived at the same final result from similar departure point. A polymorphism.

One of the most widely used models of growth applicable to the entire period of progressive postnatal ontogeny – polynomial model 1 of Preece-Baines does not always fit data from small populations that are either incomplete or show no clear adolescent growth spurt. [17, 18, 22] Although this model does not theoretically assume a priori the existence of the adolescent growth spurt it uses a defined number of parameters meant to describe adolescent growth (at least 5), values of which must be estimated irrespective of the shape of an individual’s growth curve. [35, 36] The Authors of this model could fit it sufficiently well to only 57% of British boys and 74% of British girls selected from the Harpenden Growth Study longitudinal records described by Tanner et al. [37]

Brown and Townsend when applying model 1 of Preece and Baines to longitudinal data of youths from the Yuendumu community in the Northern Territory of Australia failed to fit it to 37% of girls and i 29% boys: ”*trial curves were fitted initially to the height observations of [all children] to identify subjects whose measurements did not span the adolescent period adequately; the final selection of subjects was made after plots of the fitted curves and estimates of the biological parameters were examined”*. [22, 38] These findings indicate that growth of individuals does not always follow the “standard” of pubertal growth spurt.

The variation of human pubertal events in both their age and their magnitude indicates heterogeneity of life histories that has a significant genetic component. [6, 19, 39, 40] A complex cascade of signals in the hypothalamus that controls the onset of puberty is genetically controlled though modified by environmental conditions. [41]

Our phenotypic findings and the fact of significant genetic control of puberty indicate polymorphism of this phenomenon. This means that individuals with different genetic endowment relevant to pubertal events react differently to the same environmental stimuli that may modify their growth.

## Conclusion

Considering maladaptive character of strong pubertal growth acceleration it seems that in the less favourable for growth conditions in the past, variants of slow growth at puberty were favoured by natural selection. With the very significant relaxation of selective pressures on humans since the beginning of industrialization genotypes determining stronger growth acceleration increased in frequency. [42, 43, 44] This would be similar to relaxed natural selection increasing incidence of type 1 diabetes, [45] heritable cancers and the heritable component of obesity. [46, 47, 48] A microevolutionary change. Growth without distinct pubertal spurt is a normal polymorphic variant.

## Data Availability

All data produced in the present study are available upon reasonable request to the authors

